# Prescribing Trends of Antimicrobials in Obstetric and Gynaecological Inpatients: A Prospective Drug Utilization Study with Concurrent Antimicrobial Stewardship Audit from a Tertiary Care Hospital in Karachi, Pakistan

**DOI:** 10.64898/2026.07.16.26358229

**Authors:** Tehreem Ansari, Ale Zehra, Shazia Jabbar, Mehreen Fatima, Beenish Syed, Syed Shaukat Ali Muttaqi Shah, Amber Sabeen Ahmed, Aasma Hamid, Hafsah Ashafaq

## Abstract

**Background:** Antimicrobial resistance (AMR) disproportionately affects low- and middle-income countries (LMICs) such as Pakistan, where obstetric and gynaecological (OBGYN) patients carry high antibiotic exposure. Specialty-specific drug utilization data with concurrent stewardship audit remain scarce. This study evaluated antibiotic prescribing patterns, consumption metrics, and antimicrobial stewardship program (AMS) compliance in OBGYN inpatients at a public sector tertiary care hospital.

**Methods:** A prospective cross-sectional study was conducted in OBGYN wards of Dow University Hospital, Karachi, from 1 September to 31 October 2025. Women receiving ≥1 systemic antibiotic were included. Daily AMS rounds were conducted by an Infectious Diseases physician and pharmacist. Antibiotic consumption was measured as Defined Daily Doses (DDD) and Days of Therapy (DOT) per 1,000 patient-days (total = 821). Antibiotics were classified by WHO AWaRe (2023) framework.

**Results:** Of 812 total admissions, 278 patients (34.2%) received ≥1 antibiotic and were enrolled (205 obstetric, 73 gynaecological), generating 636 prescriptions (mean 2.29/patient). Surgical prophylaxis was the predominant documented indication (213, 33.5%); 65.1% carried no documented indication. By AWaRe classification, 53.6% were Access-group and 46.1% Watch-group. Ceftriaxone (38.4%) and metronidazole (36.8%) together represented 75.2% of prescriptions. Combined DDD/1,000 patient-days was 1,758.6 and DOT/1,000 patient-days was 1,852.7. AMS compliance was 0%.

**Conclusions:** This study documents high antibiotic prescribing burden, near-universal documentation failure, and zero AMS compliance in OBGYN inpatients at a Pakistani public sector hospital. The predominance of Watch-group antibiotics and undocumented surgical prophylaxis highlights structural stewardship gaps. Findings support urgent need for institutional OBGYN antibiotic guidelines and structured pharmacist-led AMS programs.

## Introduction

Antibiotics rank among the most frequently prescribed drugs in hospital settings worldwide, serving both prophylactic and therapeutic purposes. Their discovery transformed the management of infectious disease; however, decades of misuse in human medicine and animal husbandry have driven the emergence and global spread of antimicrobial resistance (AMR) [1]. The World Health Organization (WHO) projects that, without decisive action, AMR could cause 10 million deaths annually by 2050, surpassing cancer as a leading cause of mortality [2].

The burden falls most heavily on low- and middle-income countries (LMICs) such as Pakistan, where surveillance infrastructure is limited, antibiotic prescribing is poorly regulated, and resistance rates are among the highest recorded globally. Within hospital medicine, obstetric and gynaecological (OBGYN) patients constitute a particularly vulnerable group: antibiotics account for approximately 80% of all drugs consumed during pregnancy, and an estimated 20–40% of pregnant women receive at least one antibiotic prescription during gestation across different countries [3, 4].

In Pakistan, a multicentre point prevalence survey found that more than 60% of admitted patients were receiving antimicrobials at the time of assessment [5]. A subsequent study reported that 91% of patients admitted to gynaecology wards received one or more antimicrobials, with ceftriaxone consistently identified as the most frequently prescribed agent [5]. Longitudinal surveillance in Lahore found that 70% of participants received at least one clinically inappropriate antimicrobial, and that documentation of indication, dose, and duration was routinely absent from clinical notes[6].

The WHO AWaRe classification system — introduced in 2017 and updated in 2023 — categorises antibiotics into Access, Watch, and Reserve groups to guide stewardship prioritisation at institutional and national levels [7, 8]. Despite the availability of this framework, implementation of formal antimicrobial stewardship programmes (ASPs) in LMIC hospital settings remains limited and the evidence base for their effectiveness in this context is sparse [9, 10].

This study was conducted to address that gap. Its primary aim was to describe antibiotic prescribing patterns, quantify consumption using WHO-standard DDD and DOT metrics, compare prescribed daily doses against WHO-defined daily doses, and measure AMS team compliance in the OBGYN wards of a public sector tertiary care hospital in Karachi. A secondary aim was to characterise the distribution of AMS red flags by antibiotic prescription sequence to inform future intervention design.

## Methods

### Study design and setting

A prospective, cross-sectional observational study was conducted in the Obstetrics and Gynaecology (OBGYN) wards of Dow University Hospital (DUH), Karachi, a public sector tertiary care teaching hospital, from 1 September to 31 October 2025.

### Study population

All women admitted to the hospital in OBGYN specialty who were receiving at least one systemic antibiotic were eligible for inclusion. Women receiving non-systemic antimicrobials only (ophthalmic, otic, inhaled, or topical routes) were excluded. No minimum duration of admission was required.

### Data collection

Data were collected by a trained Infectious Diseases (ID) physician and a pharmacist using a standardized data collection proforma (Supplementary File 1) and an AMS audit form (Supplementary File 2). Information recorded included patient demographics (age, marital status), clinical data (admitting diagnosis, comorbidities, site of infection), and detailed antibiotic data for all the up to four concurrent or sequential prescriptions per patient: drug name, indication, dose, frequency, route of administration, duration, prescribing type (prophylactic, empiric, targeted, or unknown), WHO AWaRe category, ATC code, and the presence of dual coverage. Laboratory data collected included total leucocyte count, serum creatinine, creatinine clearance, liver function tests, and microbiological culture results where available. **Fig 1** provides a visual summary of the study flow, including patient admissions, enrolment criteria, data collection instruments, and daily AMS rounds.

**Fig 1.**
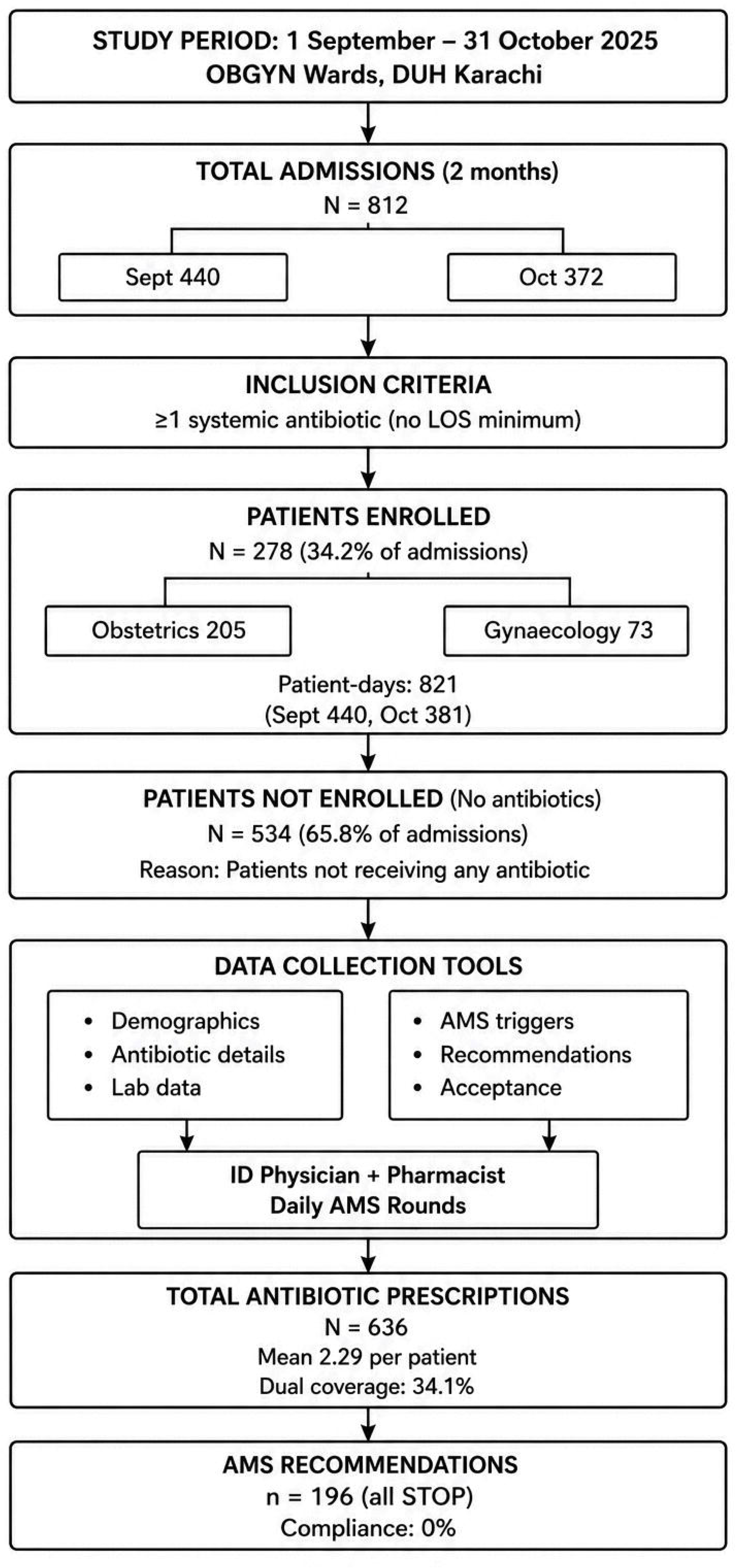
Study flow diagram: patient admissions, enrolment, data collection, and AMS outcomes. Flow diagram illustrating patient admissions to OBGYN wards over two months (September–October 2025), enrolment criteria, data collection instruments (Supplementary file 1 and 2), daily AMS rounds, and key outcomes. Of 812 total admissions, 278 patients (34.2%) received ≥1 systemic antibiotic and were enrolled. The remaining 534 admissions (65.8%) did not receive antibiotics. Enrolled patients generated 636 antibiotic prescriptions (mean 2.29 per patient). Daily AMS rounds by an ID physician and pharmacist identified 196 recommendations (all to stop antibiotics), with zero compliance (0%). DDD = defined daily dose; DOT = days of therapy.

### Data management and quality assurance

To minimize transcription errors, double data entry was performed. All the data was entered by two independent data entry operators from the paper-based proforma and AMS audit form (File S1) and transcribed into a structured Microsoft Excel database (version 16.0). The two datasets were compared to check the discrepancy. Discordant entries (n = 24, <0.5% of all fields) were resolved by confirming the original source documents and, where necessary, consultation with the ID physician and pharmacist. Any field left blank on the proforma, was coded as "not documented", illegible or ambiguous entries were coded as "unknown" after confirmation with the data collector. Range and logic checks were applied to identify out-of-range values (e.g., dose >2g for ceftriaxone or duration >30 days) and logical inconsistencies (e.g., intravenous route recorded for an oral-only antibiotic). All identified queries were verified against source documents before final dataset lock.

### WHO AWaRe Classification

Antibiotics were categorized to the WHO AWaRe (Access, Watch, and Reserve) classification system (2023). Access-agents are narrow spectrum antibiotics with lower resistance potential and are preferred choice for empiric therapy when appropriate. Watch-agents are broad-spectrum antibiotics with higher resistance potential and require closer monitoring and stewardship insight. Reserve-agents are last line antibiotics reserved for multidrug-resistant infections and should only be used when all other alternatives have failed [7].

### Antimicrobial stewardship Recommendations

AMS applicable cases were identified by daily review of patient files by an ID physician-pharmacist team. Each case was reviewed and, where applicable, a written AMS recommendation was issued and document in the patient medical record. Reasons of AMS recommendations were documented using a structured classification scheme aligned with WHO guidance (File S2). Patients discharged before the review of ID physician-pharmacist were included for drug utilisation review but excluded from the recommendation and compliance analysis. Compliance to the AMS recommendations was defined as binary: a recommendation was either accepted or not accepted by the treating team.

### Antibiotic consumption metrics

Antibiotic consumption was quantified using the Defined Daily Dose (DDD) methodology of the WHO Collaborating Centre for Drug Statistics Methodology (2023 ATC/DDD Index) [11]. DDD per 1,000 patient-days was calculated as: (total grams consumed ÷ WHO DDD) ÷ patient-days × 1,000. Days of Therapy (DOT) per 1,000 patient-days was calculated as the total number of calendar days on which a given antibiotic was prescribed, divided by patient-days and multiplied by 1,000. Total patient-days were 440 in September 2025 and 381 in October 2025 (combined: 821). The Drug Utilization 90% (DU90) segment was identified by ranking antibiotics in descending order of combined DDD/1,000 patient-days and selecting the subset accounting for 90% of total DDD volume [12].

### Prescribed daily dose analysis

The Prescribed Daily Dose (PDD) was derived for each antibiotic prescription as: PDD (g) = (single dose in mg × daily frequency) ÷ 1,000. Frequency multipliers were: once daily (OD) ×1, twice daily (BD) ×2, three times daily (TDS) ×3, four times daily (QID) ×4, and stat/pre- operative ×1. Deviation of PDD from DDD was expressed as a percentage: deviation (%) = (mean PDD/DDD − 1) × 100. Results are presented separately for September 2025, October 2025, and the combined study period.

### Statistical analysis

All statistical analyses were performed using Python version 3.9 with the following libraries: pandas (v2.1) for data cleaning, reshaping, and tabulation; NumPy (v1.26) for numerical operations; SciPy (v1.11) for all inferential statistical tests; Matplotlib (v3.8) and Seaborn (v0.13) for data visualization. All code was executed in a Jupyter Notebook environment; outputs were exported to plain text and imported into Microsoft Excel for final table formatting. No commercial statistical package was used. No imputation was performed for missing data; denominators are reported as the number of non-missing observations for each variable and are stated explicitly in each table. Continuous variables — age (years), length of stay (days), number of comorbidities, antibiotic duration (days), prescribed daily dose (g), and PDD/DDD ratio — are presented as mean ± standard deviation (SD). Where distributions were markedly right-skewed, confirmed by the Shapiro–Wilk test, median and interquartile range (IQR) or full range are additionally reported. Categorical variables are reported as counts and valid percentages calculated against the number of non-missing observations for that variable.

Each patient could receive up to four concurrent or sequential antibiotic prescriptions during a single admission, recorded as initial prescription (first antibiotics ordered) and subsequent prescriptions (second, third and fourth orders). All four prescriptions were analysed independently and in aggregate according to their order of administration. The unit of analysis for all per-antibiotic prescription calculations is the individual prescription, not the patient. For initial and subsequent prescription, the following were computed: prescription count and percentage of the total 636; AWaRe classification distribution (Access/Watch/Reserve); prescribing type distribution (Prophylactic/Empiric/Targeted/Unknown–Not Mentioned); dual antibiotic coverage rate; and mean duration (± SD). Differences in AWaRe distribution, prescribing type distribution; dual antibiotic coverage rate; and mean duration were assessed using the chi-square test or Fisher’s exact test (where expected cell counts fell below five). Differences in antibiotic duration across different prescriptions were assessed using the Kruskal– Wallis H test with Dunn’s post-hoc test and Bonferroni correction for pairwise comparisons.

The proportion of Access, Watch, and Reserve antibiotics was calculated across all 636 prescriptions and separately for each of the four prescriptions. An overall chi-square test was applied to a 3 × 4 contingency table (AWaRe category × antibiotic sequence number). A binary chi-square test (Access vs Watch+Reserve) was conducted to provide a clinically interpretable comparison. Fisher’s exact test was substituted for cells with expected frequency below five.

DDD per 1,000 patient-days was calculated as: DDD/1,000 PD = (total grams consumed ÷ WHO DDD) ÷ patient-days × 1,000. DOT per 1,000 patient-days was calculated as total calendar days of therapy divided by patient-days and multiplied by 1,000. The DU90 segment was identified by ranking antibiotics in descending order of combined DDD/1,000 PD and selecting those whose cumulative DDD volume reached 90% of the total, following Bergman et al. (1998) and Nunes et al. (2022). Monthly DOT/DDD ratios were compared descriptively [13, 14]. PDD was calculated as: PDD (g) = (single dose in mg × daily frequency) ÷ 1,000. Frequency multipliers were OD×1, BD×2, TDS×3, QID×4, stat×1. Deviation (%) = (mean PDD/DDD − 1) × 100. Results are reported separately for September, October, and the combined period. The DDD is a statistical unit for drug utilization comparison, not a recommended clinical dose; deviations are interpreted in the context of local patient characteristics and clinical indication.

For each AMS intervention, absolute frequency was computed across all prescriptions and stratified by course number (first through fourth antibiotic prescribed). AMS compliance was a binary outcome (accepted/not accepted per recommendation). The overall compliance rate was calculated as the proportion of accepted recommendations; no inferential test was applied given that compliance was uniformly zero. For comparative analysis, continuous variables, normality was assessed using the Shapiro–Wilk test. The Mann–Whitney U test (two-sided) was used for between-month comparisons of non-normally distributed variables. Chi-square or Fisher’s exact tests were applied for categorical comparisons. A two-sided α of 0·05 was used throughout. No correction for multiple comparisons was applied given the descriptive and hypothesis-generating nature of this study. The primary AMS outcome — inappropriate antibiotic prescribing — was present in 99·5% of all antibiotic prescriptions and showed virtually no variation. Logistic regression, Poisson regression, and related methods were therefore not attempted, as any such model would have yielded unstable or infinite estimates due to complete or quasi-complete separation. This limitation is acknowledged explicitly.

Binary logistic regression was performed to identify predictors of dual antibiotic coverage, defined as concurrent prescription of two or more antibiotics with overlapping spectra flagged as clinically unwarranted by the AMS team. Univariate analysis was conducted for all candidate predictors: age, department (Obstetrics vs Gynaecology), length of stay, number of comorbidities, comorbidity presence (binary), ward type, discharge before AMS round, Watch-group antibiotic use (vs Access-group), indication type (surgical prophylaxis, unknown/not mentioned, or other), and whether a culture was sent. Variables with p < 0.20 on univariate analysis were entered into a multivariate model using backward elimination with removal threshold p > 0.10. Adjusted odds ratios (aOR) with 95% confidence intervals were reported. Number of antibiotics prescribed was excluded from regression due to structural collinearity with the outcome (dual coverage requires ≥2 antibiotics). Model fit was assessed using Nagelkerke R². Statistical significance was set at p < 0.05. Analyses were performed using Python 3.x with statsmodels.

### Ethical consideration

Ethical approval was obtained from the Institutional Review Board of Dow University of Health Sciences (IRB reference: IRB-/DUHS/Approval/2024/102). Written informed consent was obtained from all participants prior to enrollment. Participants were informed of their right to withdraw from the study at any time. Data were anonymized before analysis to ensure patient confidentiality.

## Results

### Patient demographics

During the study period, there were 812 admissions to OBGYN wards (440 in September, 372 in October). Of these 278 patients (34.2%) received at least one systemic antibiotic and were enrolled in the study: 205 from Obstetrics (73.7%) and 73 from Gynaecology (26.3%). The remaining 534 admissions (65.8%) did not receive any systemic antibiotic during their hospital stay, primarily because they were admitted for normal vaginal delivery without complication, diagnostic follow-up for non-infectious conditions, elective caesarean section without risk factors, or day care admissions not requiring antimicrobial therapy.

Gynaecology patients were considerably older than their obstetric counterparts (mean 44.8 ± 15.1 years vs 28.2 ± 5.6 years), which reflects the divergent clinical caseloads of the two services. Length of stay was modestly longer in Gynaecology (mean 3.4 vs 2.7 days), consistent with a higher proportion of elective surgical admissions. Comorbidity burden was low overall but higher in the Gynaecology group (mean 0.71 vs 0.39 comorbidities per patient). Antibiotic duration was nearly identical across departments, with a mean of 2.6 days and a median of 3 days in both groups (Table 1). Of the 278 enrolled patients, 112 (40.3%) were identified as having dual antibiotic coverage, defined as concurrent prescription of two or more antibiotics with overlapping spectra deemed clinically unwarranted by the AMS team.

**Table 1.** Patient demographics and descriptive statistics by department.

| Variable | Overall (n=278) | Obstetrics (n=205) | Gynaecology (n=73) | Range |
| --- | --- | --- | --- | --- |
| Age, years (mean ± SD) | 32.6 ± 11.7 | 28.2 ± 5.6 | 44.8 ± 15.1 | 15–88 |
| Length of stay, days (mean ± SD) | 2.9 ± 1.4 | 2.7 ± 1.0 | 3.4 ± 2.1 | 0–12 |
| No. of comorbidities (mean ± SD) | 0.47 ± 0.75 | 0.39 ± 0.70 | 0.71 ± 0.86 | 0–3 |
| Antibiotic duration, days (mean ± SD) | 2.6 ± 1.1 | 2.6 ± 0.9 | 2.6 ± 1.3 | 1–9 |
| **Total prescriptions n(mean)** | **636 (2.29)** | **471 (2.30)** | **165 (2.26)** | — |

*SD = standard deviation. Antibiotic duration calculated from prescriptions with a recorded duration value (n = 202 overall, 150 Obstetrics, 52 Gynaecology, 76 excluded as 75 received antibiotic pre-op and 1 with missing duration).*

### Antibiotic prescribing patterns

The 278 patients enrolled in this study generated 636 antibiotic prescriptions in total (mean 2.29 per patient). The majority of patients received two concurrent or sequential antibiotics: 277 received an initial antibiotic prescription, 249 a subsequent prescription, 105 received a third prescription of antibiotic, and five patients received a fourth antibiotic prescription. Dual antibiotic coverage was recorded in 217 instances (34.1% of all prescriptions), rising markedly with prescription order (10.5% in initial prescriptions vs 74.3% in third-order subsequent prescriptions).

Indication was not-documented in 414 of 636 prescriptions (65.1%). Among those with a documented indication, surgical prophylaxis (SP) accounted for all 213 labelled cases. Community-acquired infections (n=2) and hospital-acquired infections (n=7) together totalled fewer than ten prescriptions. By WHO AWaRe classification (**Fig 2**), 341 prescriptions (53.6%) involved Access-group agents and 293 (46.1%) Watch-group agents; Reserve-group use was restricted to two prescriptions (0.3%). Intravenous administration accounted for 87.9% of all prescriptions (Table 2).

**Fig 2:**
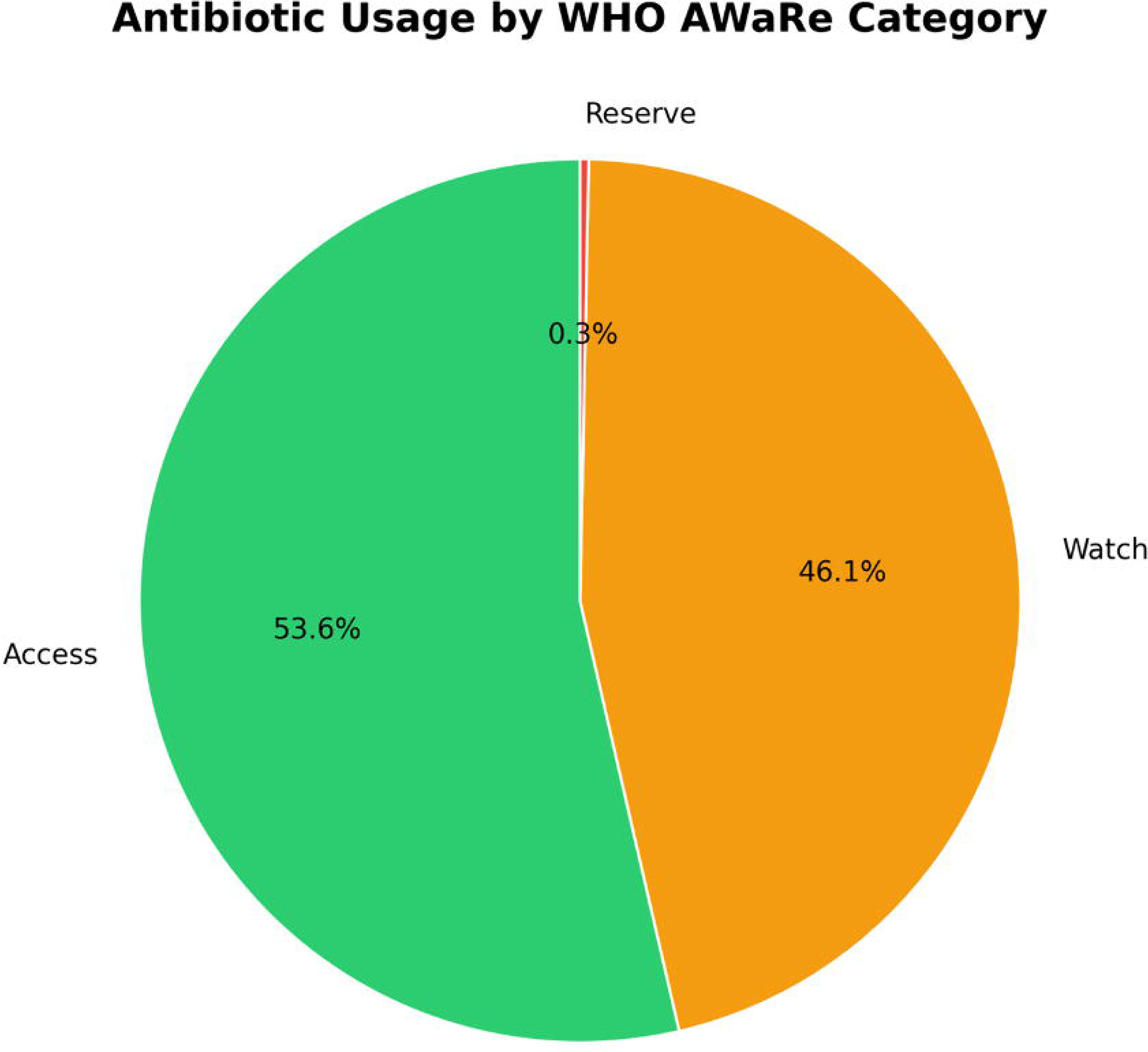
Antibiotics usage by WHO AWaRe classification. Pie chart showing the distribution of 636 antibiotic prescriptions according to the WHO AWaRe (Access, Watch, Reserve) classification system (2023). Access-group antibiotics (narrow-spectrum, lower resistance potential) accounted for 341 prescriptions (53.6%). Watch-group antibiotics (broader-spectrum, higher resistance potential) accounted for 293 prescriptions (46.1%). Reserve-group antibiotics (last-line agents for multidrug-resistant infections) were restricted to two prescriptions (0.3%).

**Table 2.** Antibiotic prescribing patterns — overall and by department.

| Parameter | Overall | Obstetrics | Gynaecology |
| --- | --- | --- | --- |
| **Total prescriptions** | **636** | **471** | **165** |
| 1st antibiotic prescription | 277 (43.6%) | 204 (43.3%) | 73 (44.2%) |
| 2nd antibiotic prescription | 249 (39.2%) | 185 (39.3%) | 64 (38.8%) |
| 3rd antibiotic prescription | 105 (16.5%) | 79 (16.8%) | 26 (15.8%) |
| 4th antibiotic prescription | 5 (0.8%) | 3 (0.6%) | 2 (1.2%) |
| **By antibiotic type** |  |  |  |
| Prophylactic | 213 (33.5%) | 157 (33.3%) | 56 (33.9%) |
| Empiric | 6 (0.9%) | 4 (0.8%) | 2 (1.2%) |
| Targeted | 3 (0.5%) | 0 | 3 (1.8%) |
| Unknown / not documented | 414 (65.1%) | 310 (65.8%) | 104 (63.0%) |
| **By indication** |  |  |  |
| Surgical prophylaxis (SP) | 213 (33.5%) | 157 (33.3%) | 56 (33.9%) |
| Community-acquired infection | 2 (0.3%) | 0 | 2 (1.2%) |
| Hospital-acquired infection | 7 (1.1%) | 4 (0.8%) | 3 (1.8%) |
| Unknown / not documented | 414 (65.1%) | 310 (65.8%) | 104 (63.0%) |
| **WHO AWaRe classification** |  |  |  |
| Access | 341 (53.6%) | 271 (57.5%) | 70 (42.4%) |
| Watch | 293 (46.1%) | 199 (42.3%) | 94 (57.0%) |
| Reserve | 2 (0.3%) | 1 (0.2%) | 1 (0.6%) |
| **Dual antibiotic coverage** | **217 (34.1%)** | **180 (38.2%)** | **37 (22.4%)** |
| Route — intravenous | 559 (87.9%) | 412 (87.5%) | 147 (89.1%) |
| Route — oral | 77 (12.1%) | 59 (12.5%) | 18 (10.9%) |

*P = Prophylactic; E = Empiric; T = Targeted; NM = Not Mentioned/documented. Percentages calculated against total prescriptions within each column. AWaRe = WHO Access/Watch/Reserve classification (2023). Dual coverage defined as concurrent prescription of two agents for anaerobic cover or documented combination therapy on the same day.*

### Antibiotic utilization by agent

Ceftriaxone and metronidazole together accounted for 75.2% of all prescriptions (n=244 and n=234 respectively), and both were administered almost exclusively by intravenous route. Amoxicillin-clavulanate ranked third (n = 106, 16.7%) and was the only widely-used agent with a meaningful proportion given orally (16.0%). Reserve-group antibiotic use was minimal: linezolid was prescribed on one occasion. Meropenem, used on three occasions, with mean treatment duration of 5.0 days, prescribed exclusively in the surgical intensive care unit (SICU). Table 3 summarises individual antibiotic utilisation data.

**Table 3.**
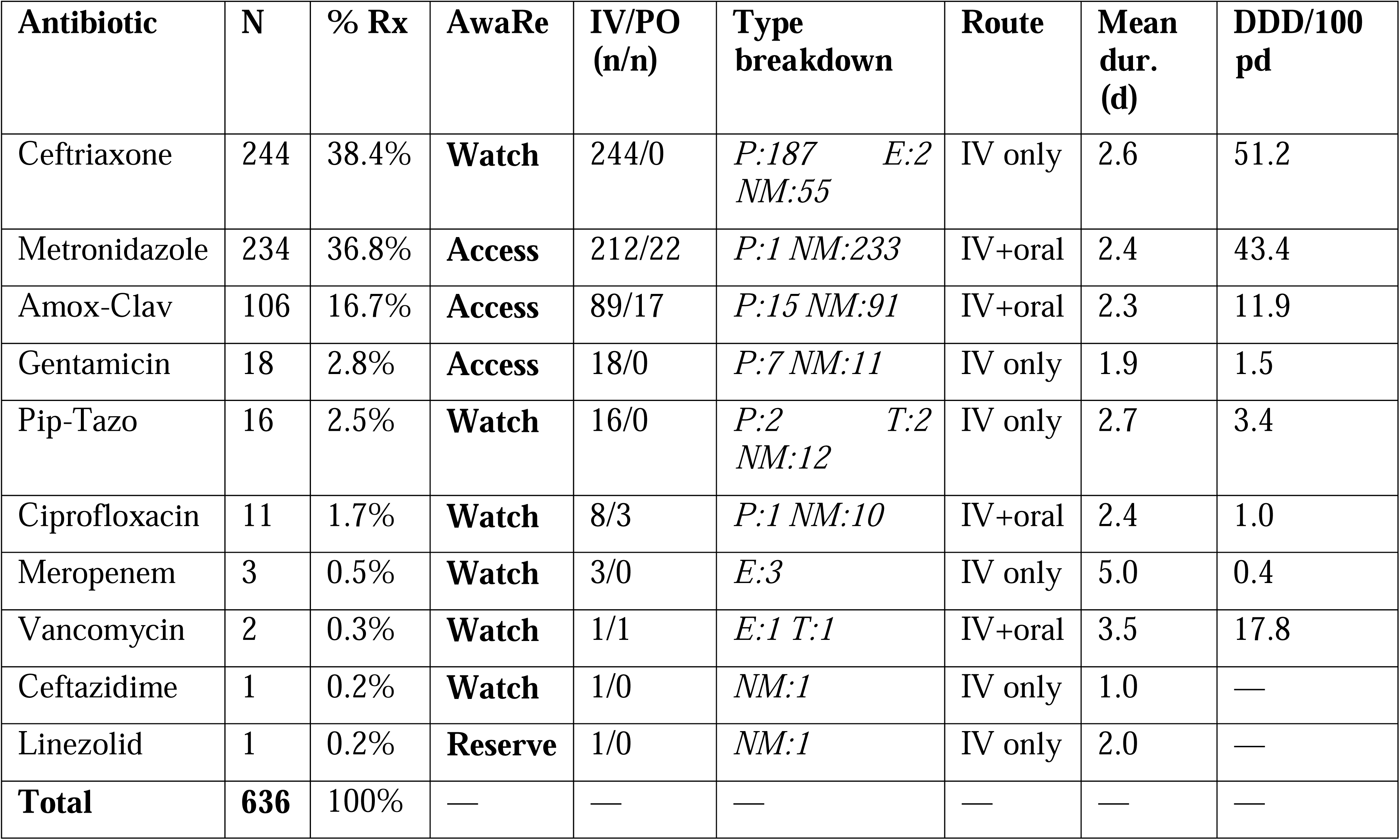
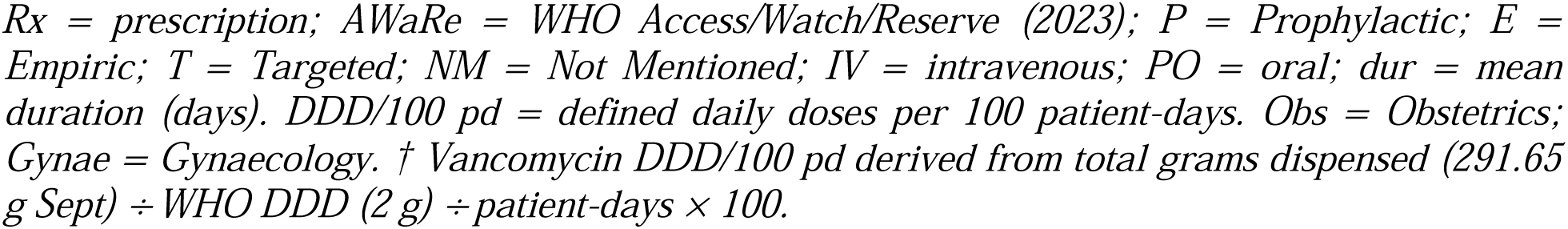
Individual antibiotic utilisation (n = 636 prescriptions, combined study period)

### Prescription distribution by ward and indication

Gynae Ward generated the highest prescription volume across all indication categories (411 of 636 total). Surgical prophylaxis prescriptions were concentrated in Gynae Ward (139) and Private Ward (55). The SICU was the only setting with a notable concentration of hospital-acquired infection (HAI) prescriptions (n = 4), consistent with the severity of illness in that unit. The high proportion of unknown/not-documented entries was uniformly distributed across all wards, reinforcing the conclusion that failure to record antibiotic indication is a system-wide rather than ward-specific problem (Fig 3).

**Fig 3:**
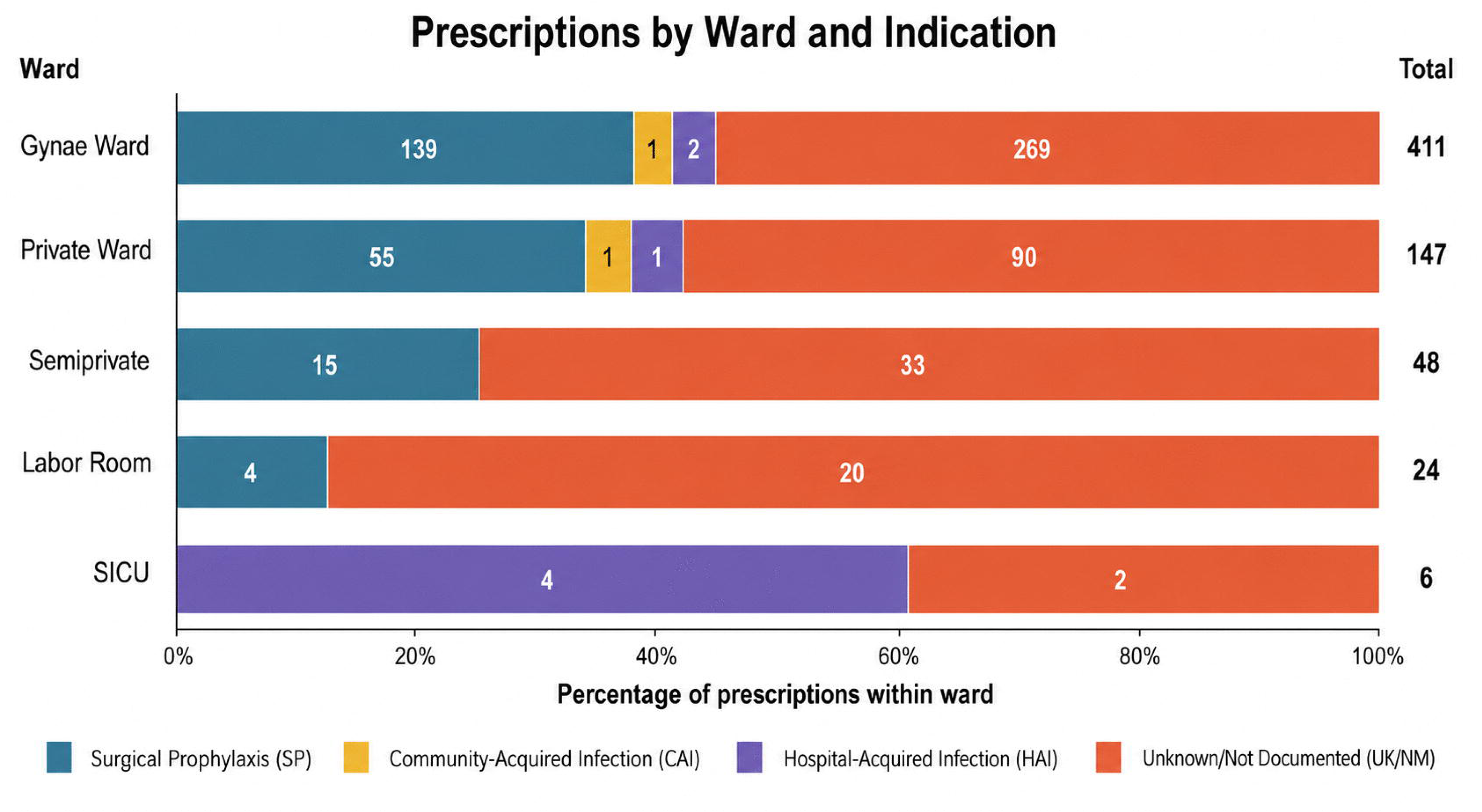
Antibiotic prescriptions by ward and indication. Bar chart showing antibiotic prescriptions (n = 636) across five OBGYN wards by indication: surgical prophylaxis (SP), community-acquired infection (CAI), hospital-acquired infection (HAI), and unknown/not documented (UK/NM). Gynae Ward had the highest prescription volume (411, 64.6%). SP was concentrated in Gynae Ward (139) and Private Ward (55). HAI prescriptions (n = 4) were confined to SICU.

Ceftriaxone and metronidazole dominated prescribing in Gynae Ward (153 and 151 prescriptions respectively) and Private Ward (61 and 55), consistent with their concurrent use as a combination surgical prophylaxis regimen. Amoxicillin-clavulanate followed a similar geographic pattern. Meropenem was prescribed almost exclusively in the SICU (2 of 3 prescriptions), and ciprofloxacin was concentrated in Gynae Ward (9 of 11 prescriptions). Piperacillin-tazobactam was used predominantly in Gynae Ward and is flagged as a stewardship priority given its Watch-group classification.

### Antibiotic consumption: DDD and DOT per 1,000 patient-days

Total combined DDD/1,000 patient-days across the study period was 1,758.6 and combined DOT/1,000 patient-days was 1,852.7. The DOT/DDD ratio was 1.59 in September and 2.15 in October, indicating a change in prescribing intensity between the two months, though total DDD/1000 PD decreased over the same period. The DU90 segment — the subset of antibiotics accounting for 90% of total DDD volume — comprised four agents: metronidazole IV (550.6 DDD/1,000 PD, 31.3% of volume), ceftriaxone (511.6, 29.1%), metronidazole oral (245.2, 13.9%), and vancomycin (177.6, 10.1%). The high DDD/1,000 PD for vancomycin reflects a small number of patients receiving prolonged prescriptions in September. **Table 4** presents combined DDD and DOT data for all agents, monthly stratified data are available in **Table S1**.

**Table 4.** Antibiotic consumption (DDD and DOT per 1,000 patient-days), with PDD/DDD ratio and DU90 segment.

| Antibiotic | ATC | AwaRe | DDD (g) | DDD /1,000 PD | DOT /1,000 PD | Mean PDD/DD ratio | DU90 |
| --- | --- | --- | --- | --- | --- | --- | --- |
| Metronidazole (IV) | J01XD01 | Access | 1.50 | 550.6 | 632.2 | 1.10 | ● |
| Ceftriaxone | J01DD04 | Watch | 2.00 | 511.6 | 627.3 | 0.84 | ● |
| Metronidazole (Oral) | J01XD01 | Access | 1.50 | 245.2 | 63.3 | - | ● |
| Vancomycin† | J01XA01 | Watch | 2.00 | 177.6 | 4.9 | 1.00 | ● |
| Amox-Clav (IV) | J01CR02 | Access | 3.00 | 170.5 | 338.6 | 1.20 | — |
| Pip-Tazo | J01CR05 | Watch | 14.00 | 32.8 | 49.9 | 0.93 | — |
| Amox-Clav (Oral) | J01CR02 | Access | 1.50 | 27.6 | 87.7 | - | — |
| Gentamicin | J01GB03 | Access | 0.24 | 20.7 | 21.9 | 1.0 | — |
| Ciprofloxacin (IV) | J01MA02 | Watch | 0.80 | 8.5 | 4.9 | 1.0 | — |
| Ciprofloxacin (Oral) | J01MA02 | Watch | 1.00 | 8.5 | 12.2 | 1.0 | — |
| Meropenem | J01DH02 | Watch | 3.00 | 4.9 | 9.7 | 0.83 | — |
| Total |  |  |  | 1,758.6 | 1,852.7 |  |  |

● DU90 = drug utilisation 90% segment. DDD = WHO defined daily dose (2023 ATC/DDD index). DOT = days of therapy. PD = patient-days (Sept 440, Oct 381, combined 821). AWaRe = WHO 2023. † Vancomycin high DDD entry driven by prolonged duration; clinical verification advised. % vol. = percentage of total combined DDD volume. Access-group DDD/1,000 PD: 952.4 (54.1%); Watch-group: 806.2 (45.8%); Reserve-group: 4.9 (0.3%). Mean PDD/DDD ratio not calculated (—) for oral metronidazole and oral amoxicillin-clavulanate due to insufficient prescription counts for reliable PDD estimation

### Prescribed daily dose versus defined daily dose

Amoxicillin-clavulanate showed the largest positive deviation from the WHO DDD, with a mean PDD/DDD ratio of 1.20 (+20.4% deviation), reflecting frequent prescribing at 1.2 g three times daily (3.6 g/day) against the WHO IV DDD of 3.0 g. Ceftriaxone showed a consistent negative deviation of −16.1% overall (mean PDD/DDD ratio 0.84), attributable to predominant use at 1 g per dose (single or twice-daily dosing) where the WHO DDD is 2 g. Metronidazole, gentamicin, and ciprofloxacin were prescribed at doses closely concordant with their respective WHO DDDs. Full PDD-versus-DDD statistics, including monthly stratification, are presented in **Table S2**.

### AMS applicable cases, interventions and compliance

A total of 196 AMS applicable cases were identified and written recommendations/interventions were proposed during the study period. Compliance with AMS team recommendations was 0% — no recommendation was accepted by treating teams during the study period. Five categories of AMS applicable cases were identified: unnecessary broad-spectrum antibiotic selection (n = 96), overprescribing through excessive dose or duration (n = 80), inappropriate drug selection (n = 66), dual anaerobic coverage without clear clinical justification (n = 65), and extended surgical antibiotic prophylaxis beyond the single-dose standard (n = 52) (Fig 4). A total 46 patients were discharged before AMS rounds were carried out, hence recommendations could not be given for these cases.

**Fig 4.**
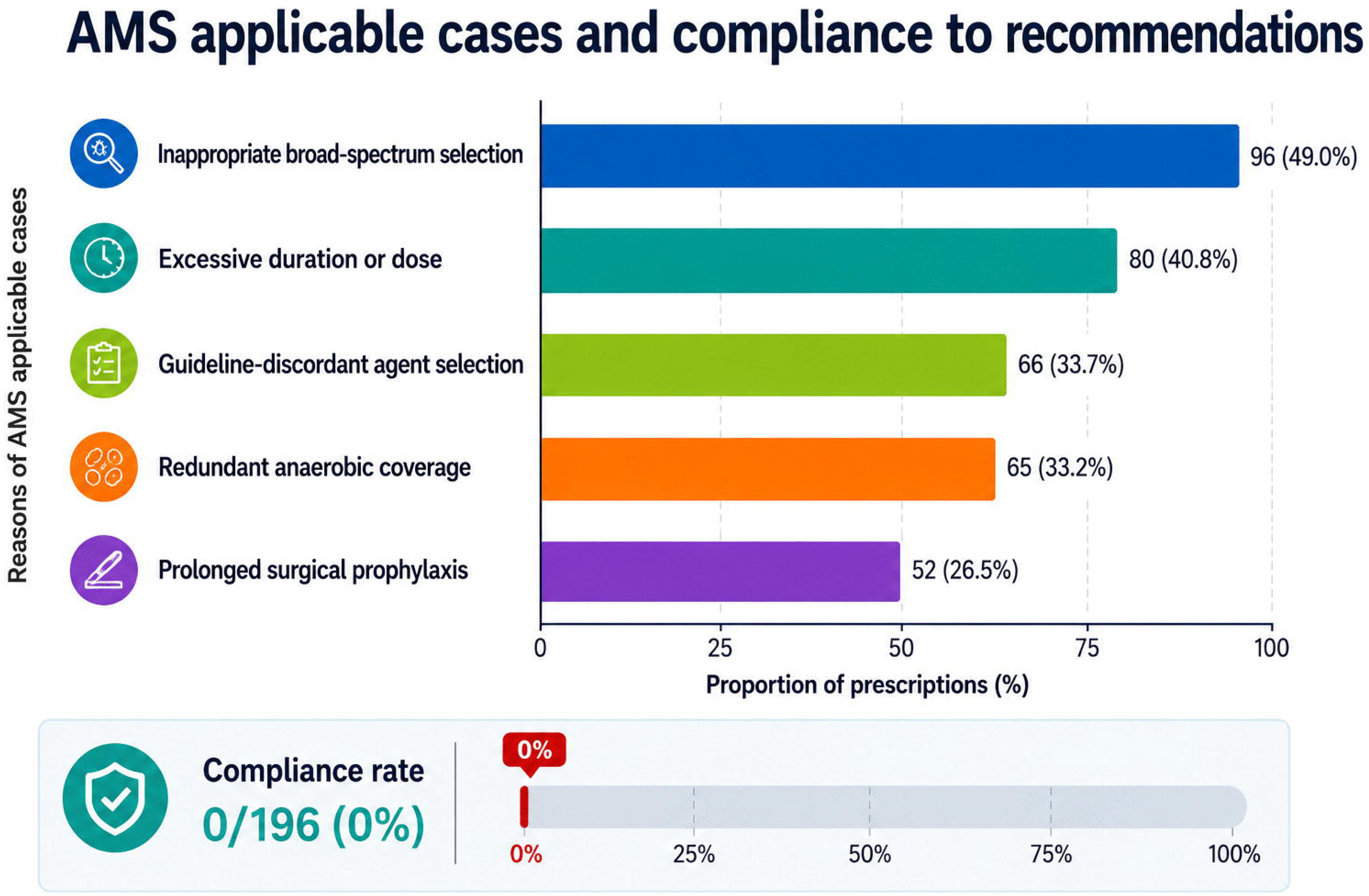
AMS intervention frequencies and compliance (n= 196 recommendations) Bar chart showing the frequency (n and %) of five AMS intervention categories. Percentages sum to >100% due to multiple triggers per prescription.

### Predictors of dual antibiotic coverage

Multivariate logistic regression was performed to identify independent predictors of dual antibiotic coverage (Table 5). After backward elimination, three variables remained in the final model (χ² = 46.27, df = 3, p < 0.001; Nagelkerke R² = 0.123). Gynaecology patients had significantly lower adjusted odds of dual coverage compared to Obstetrics patients (aOR 0.42, 95% CI 0.22–0.80, p = 0.008), representing a 58% reduction in odds. Patients receiving Watch-group antibiotics also had substantially lower odds of dual coverage compared to those receiving Access-group antibiotics (aOR 0.08, 95% CI 0.03–0.22, p < 0.001), a 92% reduction. Discharge before AMS round showed a borderline association with lower odds of dual coverage (aOR 0.54, 95% CI 0.28–1.04, p = 0.064), suggesting possible under-ascertainment in patients not formally reviewed. Age, length of stay, comorbidity burden, and study month were not significant predictors in either univariate or multivariate analysis.

**Table 5.** Logistic regression analysis: predictors of dual antibiotic coverage.

| Variable | OR (95% CI) | p-value | aOR (95% CI) | p-value |
| --- | --- | --- | --- | --- |
| Age (years) | 0.98 (0.96–1.00) | 0.102 | — | — |
| Gynaecology (vs Obstetrics) | 0.46 (0.25–0.82) | 0.009 | 0.42 (0.22–0.80) | 0.008 |
| Length of stay (days) | 1.00 (0.84–1.19) | 0.979 | — | — |
| Number of comorbidities | 0.80 (0.57–1.11) | 0.186 | — | — |
| Comorbidity present (yes vs no) | 0.81 (0.49–1.35) | 0.430 | — | — |
| Private ward (vs Gynae ward) | 0.84 (0.47–1.50) | 0.566 | — | — |
| Discharged before AMS round | 0.67 (0.38–1.19) | 0.176 | 0.54 (0.28–1.04) | 0.064† |
| Watch-group antibiotic (vs Access) | 0.24 (0.13–0.48) | <0.001 | 0.08 (0.03–0.22) | <0.001 |
| Indication: Surgical prophylaxis | 1.10 (0.65–1.87) | 0.710 | — | — |
| Indication: Unknown/not mentioned | 1.28 (0.74–2.21) | 0.384 | — | — |
| Culture sent (yes vs no) | 1.04 (0.29–3.78) | 0.948 | — | — |

OR = unadjusted odds ratio; aOR = adjusted odds ratio. Dependent variable: dual antibiotic coverage (present in 112/278, 40.3%). N = 278; number of events = 112. Reference categories: Obstetrics department, Access-group antibiotic, Gynaecology ward, not discharged before AMS round. Multivariate model included variables with $p < 0.20$ (gynaecology, Watch-group, discharged before AMS round, number of comorbidities). Number of comorbidities removed during backward elimination ( $p = 0.727$ ). Model statistics: $\chi^2 = 46.27$ ( $df = 3$ ), $p < 0.001$ ; Nagelkerke $R^2 = 0.123$ . † $p < 0.10$ (trend)

## Discussion

This prospective cross-sectional study of antimicrobial prescribing patterns in OBGYN inpatients provide several important insights. To our knowledge, this is the first study from Pakistani public sector tertiary care hospital to provide comprehensive assessment of antimicrobial utilization in an OBGYN department, using WHO-standardized metrics (DDD, DOT, and AWaRe classification), and PDD comparisons. While previous PPS have described antimicrobial use across mixed hospital populations [15, 16], and A-PLUS trial reported intrapartum and postpartum antibiotic use across seven LMICs [17], the present investigation offers three distinct contributions. First, it provides ward-specific analysis, revealing differential prescribing patterns between Obstetrics and Gynecological patients. Second, it quantifies antibiotic consumption using both DDD and DOT metrics, enabling direct comparison with international surveillance data. Third, it identifies systematic dosing anomalies and potential AMS intervention triggers through structured ID physician-pharmacist review, providing an evidence-informed roadmap for future AMS initiatives in this setting.

In the present study, the antibiotic prevalence in OBGYN wards was 34.2% of all admissions (278/812). This is lower than prevalence reported from mixed ward PPS in similar settings, which range from 57.6% in India to 83.3% at the same institution [16, 18]. The multicentre PPS conducted by Saleem et al. across 13 hospitals in Punjab, Pakistan, reported a high percentage of hospitalized patients received at least one antibiotic [19]. The lower overall prevalence observed in present OBDYN cohort is expected, as substantial proportion of admissions do not require antimicrobial therapy. The predominance of ceftriaxone as the most frequently prescribed antimicrobial (38.4%) aligns with findings from multiple LMIC settings. Zehra et al. from the same institution, reported ceftriaxone as most prescribing antibiotic in general hospital population with 18.5% of prescription, less than half the proportion observed in the present OBGYN cohort [16]. Saleem et al. reported 37.2% of all prescriptions of ceftriaxone, in Pakistani hospitals [20]. The observational study from Ethiopia by Gashaw et al. reported ceftriaxone as the most frequently used agent, with a utilization rate of 44.65 DOTs per 100 patient-days [21]. Similarly Indian multicentre survey reported ceftriaxone, accounting for 14.9% of all antibiotics (highest proportion) [18]. The consistent predominance of ceftriaxone (Watch-agent) suggests a pattern of empirical broad-spectrum prescribing, not limited to geographical boundaries, requiring urgent stewardship attention. This predominance of ceftriaxone is a divergence from established national and regional recommendations for OBGYN surgical prophylaxis. The National Action Plan (NAP) of Pakistan (2023) recommends cefazolin as the preferred agent for SP in OBGYN procedures [22]. Similarly, guidelines from other LMICs, including Indonesian Ministry of Health, endorse cefazolin, as first line agent for most uncomplicated OBGYN surgeries [23]. In the present study, cefazolin was not prescribed on a single occasion, instead ceftriaxone accounted for 38.4% of all prescriptions, almost exclusively for SP. This complete absence of the guideline recommended Access-group alternative represents a significant stewardship gap, suggesting that institutional prescribing are not aligned with standards. This substitution of ceftriaxone to cefazolin may be associated with factors like dosing convenience, perceived broader spectrum of cefazolin, lack of formulary restrictions, or absence of local guidelines. The combination of ceftriaxone with metronidazole (75.2% of prescriptions) in the present study, appears to represent a de facto standard for SP in this setting. While this combination has an evidence base in gynaecological surgery, the WHO AWaRe framework and international OBGYN AMS guidelines increasingly favor amoxicillin-clavulanate or cefazolin plus metronidazole (both Access-group agents) for most uncomplicated procedures. Implementing these recommendations for SP would reduce Watch-group exposure and improve meeting stewardship targets.

The proportion of Access-agents in this study (53.6%) was below the WHO-recommended threshold 60% of total antibiotic consumption at the hospital level [24]. Other studies have reported variable Access proportions: 32.67% in India, while 48.3% in an Ethiopian study [18, 21]. The proportion of use Watch-agents in the current study (46.1%) is broadly comparable to Ethiopian study (51.7%), but higher than the proportion observed in high-income settings, where Access-agents are prevalent in the majority of inpatient settings [21]. Zehra et al. reported a higher Watch category (80.8%) in the same institution, with the different case mix (55.5% CAIs patients) compared to the present OBGYN cohort [16]. The finding that 65.1% of prescriptions lacked any documented indication, exceeds rates reported in other Pakistani studies (34.2% -46.9%) [15, 25]. The study from the same institution classified 96% of prescriptions, suggesting the documentation deficit in the present study may be associated specifically to OBGYN wards [16]. This gap may be associated with differences in documentation practices, data collection methodologies, or the specific OBGYN clinical contexts.

Prior studies in Pakistan, describing prescribing patterns (frequencies and proportions), and using WHO PPS surveys, represent existing PPS literature, to which the dosing analysis in this study offers methodological extension [15, 16, 26]. When interpreting the PDD/DDD deviations, it is important to recognize that the WHO DDD and DOT are statistical units for drug consumption measurement, not a clinical dosing recommendation, and variation in consumption not necessarily indicate inappropriate use [27]. In this study, appropriateness was assessed using documented indication, adherence to the standards, AWaRe classification, and AMS opportunities identified. High consumption of vancomycin (1.776 DDD/1000PD) warrants clinical review, but does not signify the inappropriateness, while high ceftriaxone consumption (511.6 DDD/1000PD) raises concern given that 76.6% of prescriptions were SP, where Access-group alternatives are recommended. Ceftriaxone prescribed at a mean daily dose of 1.68 g, with a deviation of (-16.1%) from standard DDD, represents a novel finding not previously reported in the Pakistani OBGYN literature. The use of 1g dose without weight-based or indication-specific adjustment reflects a lack of standardized prescribing protocols, a stewardship concern, though not necessarily constitute sub-therapeutic dosing for surgical prophylaxis. 1 g single-dose of ceftriaxone has been used as SP in OBGYN [28], although major guidelines recommend first-generation cephalosporins such as cefazolin as the preferred agents [29]. The positive PDD/DDD deviation observed for amoxicillin-clavulanate (+20.4%) can be associated partly with the WHO DDD benchmark (3.0 g IV) compared to common clinical practice at 1.2 g three times daily (3.6 g/day) [30]. Metronidazole, mean daily dose of 1.61 g, modestly exceeding the WHO DDD of 1.5 g with a deviation of (4.8%). While the clinical significance of these small deviation is uncertain, it indicates a lack of dose standardization and suggests the need for structured local prescribing guidelines.

The zero AMS compliance rate to the daily ID physician-pharmacist recommendations is particularly instructive, demonstrating that voluntary, recommendation-only AMS interventions without organizational mandate was ineffective. This finding is consistent with evidence from LMIC settings showing that persuasive interventions alone are often insufficient [31]. In resource-limited settings, the absence of local antibiotic guidelines, no restrictions of formulary, and lack of managerial mandate does make stewardship recommendations ineffective [32]. Systematic reviews of AMS programs in LMICs indicate that, successful AMS programs involves combination of enabling elements (guideline), persuasive elements (audit and feedback), and structural or restrictive components [33]. The failure in the present study may be attributed to the absence of enabling elements like, absence of, local guidelines, pharmacist follow-up, or managerial accountability, rather than voluntary nature of recommendations.

In this study, the antibiotics were used as SP for almost all the prescriptions with documented indication (33.5% of total prescriptions), this finding is consistent with 82.3%–97.7% of prophylactic antibiotic use in the A-PLUS trail [17]. While in a previous study from hospital in Pakistan for gynecological surgeries, adherence to the guidelines was 40.7%, timing adherence of 56.4%, with ceftriaxone (22.4%) and cefazolin (22%) commonly prescribed agents [34]. In the present study, the 65.1% undocumented indication should not be interpreted as a documentation lapse but rather a structural deficiency, indicating: that the basic data required for prescribing appropriateness assessment are systematically absent, making any meaningful AMS evaluation impossible [35]. The prevalence of inappropriate broad-spectrum selection and prolonged SP, identifies a clear stewardship target, as systemic reviews have demonstrated that SP >24 hours provides no additional benefit over single-dose regimens [36, 37]. These gaps suggest that effective AMS in this setting requires transition from voluntary and educational approach to structural interventions, including mandatory indication fields, Watch-group restriction, automatic stop-orders, and pharmacist-led audit along with prescribers accountability [38, 39].

The findings have several implications for AMS in LMIC OBGYN settings. First, local Obstetrics and Gynecological antimicrobial prescribing guidelines should be developed to establish indications, preferred Access-category agents in SP, standardized dosing, and treatment durations. Second, the 65.1% undocumented indication rate requires mandatory indication fields accompanied with regular audit and feedback. Third, ceftriaxone to cefazolin switchover in the OBGYN settings to improve the Access-Watch ratio. Fourth, potential AMS triggers inform targeted initiatives. The WHO has explicitly called for the implementation of AMS programs in LMICs, based on the local baseline data to inform context specific interventions [40]. The present study directly addresses this call by providing granular, ward-specific baseline data to guide stewardship activities in a Pakistani OBGYN setting. Furthermore, the methodological approach employed, incorporating WHO metrics (DDD, DOT, AWaRe classification) alongside structured AMS review, offers a replicable model for similar settings in other LMIC hospitals.

In this study, Gynaecology patients had 58% lower adjusted odds of dual coverage compared to obstetric patients, this finding reflects routine clinical practice in the obstetric ward of co-prescribing metronidazole with amoxicillin-clavulanate following lower segment caesarean section (LSCS), a dual anaerobic coverage regimen that is not supported by evidence. Patients receiving Watch-group antibiotics had 92% lower adjusted odds of dual coverage, indicating that dual coverage is predominantly an Access-group prescribing problem, driven by the combination of Access agents (metronidazole + amoxicillin-clavulanate) rather than Watch-group agents like ceftriaxone. The finding that dual coverage was an Access-group prescribing problem is consistent with previous studies from LMIC settings [41]. Similar patterns of redundant anaerobic coverage have been documented in surgical wards in Kenya and Ghana, where co-prescription of metronidazole with beta-lactam antibiotics accounted for a substantial proportion of inappropriate prescribing [42, 43]. The borderline association for discharge before AMS round, likely reflects ascertainment bias, as patients discharged early were not formally reviewed, potentially leading to under-identification of dual coverage in this subgroup. Age, length of stay, comorbidity burden, and study month were not significant predictors, suggesting that dual coverage is driven by institutional prescribing norms rather than patient-specific clinical factors, a finding consistent with qualitative studies from South Asia highlighting the role of habit and perceived convenience in antibiotic prescribing decisions [44]. These findings identify a clear, actionable stewardship target: replacement of routine dual anaerobic coverage post-LSCS with a standardized single-agent prophylaxis protocol, coupled with early daily AMS rounds to review all patients before discharge.

The principal strengths of this study include: the prospective design, with systematic data collection conducted by a trained ID physician and pharmacist, minimizing recall and ascertainment biases; the use of standardized WHO antibiotic consumption (DDD, DOT) metrics and AWaRe classification enables direct comparison with national and international data; the separate analysis of Obstetrics and Gynaecology wards provides prescribing patterns between these two patient populations; the identification of potential AMS intervention offers an evidence based roadmap for future AMS. Despite these strengths, several limitations must be acknowledged. First, this study was conducted at a single institution, limiting the generalizability to other settings. Second, the sample size (n = 278), while adequate for descriptive purposes, is smaller compared to multicenter surveys in the regions[17]. Third, the two-month data collection period (September – October 2025) may not capture seasonal variations in infection prevalence or antimicrobial prescribing practices. Fourth, the high proportion of undocumented indications restrict the complete classification of prescriptions, limiting the assessment of prescribing appropriateness. Fifth, the study didn’t collect detailed clinical data on the 534 admissions who did not receive antibiotics, limiting direct comparison between antibiotic treated and untreated patients. Sixth, the unavailability of microbiological culture and sensitivity testing data for the majority of patients, restricts any assessment of the appropriateness of empirical antimicrobial selection. Seventh, the WHO DDD methodology, has recognized limitations in AMS application in OBGYN and surgical prophylaxis, where dosing may deviate from standard DDD values based on patient weight, renal function, or procedural characteristics [27]. Finally, the logistic regression model explained only 12% of the variance (Nagelkerke R² = 0.123), indicating that unmeasured factors beyond the included predictors contribute to dual antibiotic coverage.

The data presented in this study establish a foundation for future initiatives, including a multi-centre PPS across Pakistani OBGYN units, a qualitative study of prescriber attitudes, a prospective pre-post AMS intervention study, ceftriaxone-to-cefazolin substitution, standardized dosing protocols, and pharmacist-led interventions and audit to provide the compliance and outcome data not captured in the present investigation. Focus group discussions or surveys are needed to explore surgeons’ barriers and reluctance in accepting AMS recommendations, along with improved communication strategies. Future studies should also assess the impact of these interventions on surgical site infection rates.

## Conclusion

This prospective drug utilization study of OBGYN inpatients at a Pakistani public sector tertiary care hospital documents a high antibiotic prescribing burden (34.2% of admissions), near-universal documentation failure (65.1% of prescriptions without documented indication), and zero AMS compliance (0%). The predominance of Watch-group antibiotics (46.1%), particularly ceftriaxone and metronidazole (75.2% of all prescriptions), and the complete absence of the guideline-recommended Access-group agent cefazolin for surgical prophylaxis represent significant AMS gaps. The zero compliance rate to the recommendations demonstrates that voluntary, recommendation-only AMS interventions are ineffective without organizational mandate, structured follow-up, and enabling elements such as local antimicrobial guidelines and managerial accountability.

These findings support an urgent need for context-specific interventions. Institutional OBGYN antibiotic guidelines prioritizing Access-group agents for SP with single-dose protocols are required. Mandatory indication fields with regular audit and feedback must address the documentation deficit. Transition from voluntary to structural AMS interventions (including formulary restriction, automatic stop-orders, and pharmacist-led audit with prescriber accountability) is essential. The methodological framework employed offers a replicable model for similar needs assessments in other LMIC hospital settings.

## Data Availability

All data produced in the present study are available upon reasonable request to the authors

## Abbreviations

AMS: Antimicrobial Stewardship Program
ATC: Anatomical Therapeutic Chemical AWaRe: Access, Watch, Reserve
DDD: Defined Daily Dose
DOT: Days of Therapy
ID: Infectious Diseases
IRB: Institutional Review Board
LMIC: Low- and Middle-Income Country
OBGYN: Obstetrics and Gynaecology
PDD: Prescribed Daily Dose
PPS: Point Prevalence Survey
WHO: World Health Organization

## Acknowledgements

The authors acknowledge Prof. Dr. Jahan Ara Hassan, Pro Vice Chancellor and Head of Department, Obstetrics and Gynecology, Dow University of Health Sciences (DUHS), for her invaluable on-site mentorship and support.

## Author Contributions

TA: Conceptualisation, methodology, data collection, AMS rounds, writing – original draft, corresponding author; AZ: Conceptualisation, methodology, data curation, statistical analysis, writing – review & editing; SJ: Data collection, clinical oversight (OBGYN), validation, writing – review & editing; MF: AMS rounds, data collection, writing – review & editing; BS: AMS rounds, data collection, writing – review & editing; SSAMS: Formal analysis, data visualisation, writing – review & editing; ASA: Data interpretation, writing – review & editing; AH: Data collection, pharmacy services coordination, writing – review & editing; HA: Data collection, pharmacy services coordination, writing – review & editing.

## Data Availability Statement

The anonymised dataset supporting the conclusions of this article is available from the corresponding author upon reasonable request.

