## Supplementary material for "Prescribing Trends of Antimicrobials in Obstetric and Gynaecological Inpatients: A Prospective Drug Utilization Study with Concurrent Antimicrobial Stewardship Audit from a Tertiary Care Hospital in Karachi, Pakistan": File S1

**Data Collection Tool**

**Prescribing trends of antimicrobials in Gynecological and Obstetrical patients: A call for Antimicrobial Stewardship**

Participant’s study number __________________

**Section 1: Demographics:**

1. Patient ID (MR number):
2. Contact number:
3. Age/Date of Birth:
4. Marital status: Single (	)	Married (	)	Divorced (	)
5. Date of Admission:
6. Date of Discharge:
7. Location:
8. Admitting diagnosis:
9. Type of Patient: Gynaecological

Obstetric (if Obs, specify gestational age in weeks) _______

**Section 2: Clinical information**

10. Co- morbid conditions (specify):
11. Site of Infection (if any)
    A. Central line
    B. CNS
    C. Respiratory
    D. CVS
    E. Intraabdominal/GI
    F. Genital tract
    G. Urinary tract
    H. Bone and joint
    I. SSTI
    J. SSI
    K. Unknown/unidentified____________________
12. Surgery performed during this procedure: Yes (if yes, proceed to next question)	No
    12.1. Name of procedure	Date of procedure

**Section 4: Lab information**

13. Creatinine
14. Creatinine Clearance
15. LFTs (Recent most)
    15.1. Total Bilirubin___ Direct Bilirubin______SGPT_____SGOT_____ALP____GGT_____
16. Culture sent?	No	Yes (if yes, proceed to next question)
    16.1. Site ((line, peripheral blood, sputum, bronchial wash, tracheal aspirate, Fluid-CSF/ ascites/pleural/other, tissue, pus, HVS, urine)
    16.2. Date of sample collection
    16.3. Name of organism(s) identified

**Section 4: Medication Information:**

| **Name of Antibiotic** |
| --- |
| **Start date** |
| **End date** |
| **Duration (days)** |
| **Dose (in mg or IU)** |
| **Frequency** |
| **Route (PO,IV,IM)** |
| **Type (Empirical, targeted, prophylaxis, pre-emptive)*** |
| **Indication (e.g. HAI, CAI)** |
| ***incase of prophylaxis, please mention surgical or medical** |

**Section 5: Information about Antimicrobial stewardship (AMS)**

1. Antimicrobial stewardship applicable? Yes (if yes, proceed to next question)	No
2. Reason for AMS (anti-microbial stewardship):
   A. Overprescribing
   B. Inappropriate drug selection
   C. Unnecessary broad spectrum antibiotic
   D. Inappropriate dose
   E. Inappropriate frequency/interval
   F. Inappropriate route
   G. Extended duration
   H. Drug-bug mismatch
   I. Use of restricted/ watched antimicrobial
   J. Dual coverage
   K. IV to oral switch
   L. Extended SAP
   M. Escalation of antimicrobials required

| Name of  Antimicrobial | Reason for AMS  (please select all  from above, where applicable) | AMS/ID team recommendation (s) | Compliance with  recommendation(yes/No) |
| --- | --- | --- | --- |
