## Supplementary material for "Prescribing Trends of Antimicrobials in Obstetric and Gynaecological Inpatients: A Prospective Drug Utilization Study with Concurrent Antimicrobial Stewardship Audit from a Tertiary Care Hospital in Karachi, Pakistan": File S2

ANTIMICROBIAL STEWARDSHIP FORM

Date of AMS________

| | | |
|---|---|---|
| MR number | | Name |
| Age | | Gender |
| Neonate | Full term | Premature |
| Weight | | Height/length |
| Date of Admission | | Location |
| Specialty | | Diagnosis |
| Physician's name | | Allergy |
| Creatinine | | Creatinine Clearance |
| LFTs (Recent most) | TB______ DB______ SGPT______ SGOT______ ALP_____ GGT______ | |

Brief History (*use additional paper for more history*)

### Antimicrobial Prescription Details

| S.No | Antimicrobial Agent(s) | Indication | Date of initiation | Date of stop | Route | *Frequency* | *Duration in days* |
|---|---|---|---|---|---|---|---|

**AMS Intervention & Recommendations**

**Pharmacist Intervention: Yes/No**

______________________________________________

______________________________________________

| S.No | Reason for Intervention (*Select all that apply*) | Name of antimicrobial(s) |
|---|---|---|
| 1 | Overprescribing | |
| 2 | Inappropriate drug selection | |
| 3 | Unnecessary broad spectrum antibiotic | |
| 4 | Inappropriate dose | |
| 5 | Inappropriate frequency/interval | |
| 6 | Inappropriate route | |
| 7 | Extended duration | |
| 8 | Extended SAP | |
| 9 | Drug-bug mismatch | |
| 10 | Use of restricted/ watched antimicrobial | |
| 11 | Dual coverage | |
| 12 | IV to oral switch | |
| 13 | Other | |

| Name of Antimicrobial Agent | Recommendation |
|---|---|

**Reviewer Name and signature**______________________________

**Prescriber's Agreement with Recommendations** YES/NO (if no, please provide rationale)

---------------------------------------------------------------

---------------------------------------------------------------

---------------------------------------------------------------

**Compliance with Stewardship Recommendations** Yes No Unspecified

CS CamScanner
