## Supplementary material for "Prescribing Trends of Antimicrobials in Obstetric and Gynaecological Inpatients: A Prospective Drug Utilization Study with Concurrent Antimicrobial Stewardship Audit from a Tertiary Care Hospital in Karachi, Pakistan": Table S1

**Monthly antibiotic consumption (DDD and DOT per 1,000 patient-days), September vs October 2025**

| **Antibiotic** | **Sept DDD/1,000 PD** | **Oct DDD/1,000 PD** | **Sept DOT/1,000 PD** | **Oct DOT/1,000 PD** |
| --- | --- | --- | --- | --- |
| Metronidazole IV | 531.8 | 572.2 | 611.4 | 656.2 |
| Ceftriaxone | 483.0 | 544.6 | 534.1 | 734.9 |
| Metronidazole oral | 442.4 | 17.5 | 59.1 | 68.2 |
| Vancomycin | 331.4 | 0.0 | 9.1 | 0.0 |
| Amoxicillin-clavulanate IV | 51.8 | 307.6 | 177.3 | 524.9 |
| Amoxicillin-clavulanate oral | 36.4 | 17.5 | 81.8 | 94.5 |
| Piperacillin-tazobactam | 24.1 | 42.9 | 52.3 | 47.2 |
| Gentamicin | 27.3 | 13.1 | 27.3 | 15.8 |
| Ciprofloxacin IV | 15.9 | 0.0 | 9.1 | 0.0 |
| Ciprofloxacin oral | 4.6 | 13.1 | 11.4 | 13.1 |
| Meropenem | 9.1 | 0.0 | 18.2 | 0.0 |
| **Total patient-days** | **440** | **381** | **440** | **381** |

**DDD = defined daily dose. DOT = days of therapy. PD = patient-days. September 2025 patient-days = 440; October 2025 patient-days = 381. Vancomycin DDD/1,000 PD of 331.4 in September reflects a small number of patients receiving prolonged courses; this finding warrants clinical review.**
