## Supplementary material for "Prescribing Trends of Antimicrobials in Obstetric and Gynaecological Inpatients: A Prospective Drug Utilization Study with Concurrent Antimicrobial Stewardship Audit from a Tertiary Care Hospital in Karachi, Pakistan": Table S2

**Prescribed daily dose (PDD) vs defined daily dose (DDD): descriptive statistics and monthly deviation (modelled on Nunes et al., Front. Pharmacol. 2022)**

|  | **DDD (g)** | **September 2025 (PD = 440)** | | | **October 2025 (PD = 381)** | | | **Combined (PD = 821)** | | |
| --- | --- | --- | --- | --- | --- | --- | --- | --- | --- | --- |
| **Antibiotic** |  | **n** | **Mean PDD ± SD (g)** | **Ratio ± SD** | **n** | **Mean PDD ± SD (g)** | **Ratio ± SD** | **Dev.** | **n** | **Deviation from DDD** |
| Ceftriaxone | 2.00 | 108 | 1.689 ± 0.471 | 0.844 ± — | 102 | 1.667 ± 0.474 | 0.833 ± — | **-16.7%** | 210 | **-16.1%** |
| Metronidazole | 1.50 | 108 | 1.649 ± 0.637 | 1.099 ± — | 95 | 1.484 ± 0.067 | 0.989 ± — | -1.1% | 203 | +4.8% |
| Amox-Clav* | 2.75 | 47 | 3.191 ± 0.705 | 1.234 ± — | 44 | 3.382 ± 0.631 | 1.173 ± — | **+17.3%** | 91 | **+20.4%** |
| Pip-Tazo | 14.00 | 7 | 12.536 ± 2.551 | 0.895 ± — | 7 | 13.500 ± 0.000 | 0.964 ± — | -3.6% | 14 | **-7.0%** |
| Gentamicin | 0.24 | 7 | 0.240 ± 0.000 | 1.000 ± — | 4 | 0.240 ± 0.000 | 1.000 ± — | 0.0% | 11 | 0.0% |
| Meropenem‡ | 3.00 | 2 | 2.500 ± 0.707 | 0.833 ± — | — | — | — | — | 2 | **-16.7%** |
| Ciprofloxacin* | 0.90 | 4 | 0.850 ± 0.100 | 1.000 ± — | 2 | 1.000 ± 0.000 | 1.000 ± — | 0.0% | 6 | 0.0% |
| Vancomycin‡ | 2.00 | 1 | 2.000 ± — | 1.000 ± — | — | — | — | — | 1 | 0.0% |
| Linezolid‡ | 1.20 | — | — | — | 1 | 1.200 ± — | 1.000 ± — | 0.0% | 1 | 0.0% |

*PDD = prescribed daily dose (single dose × daily frequency, in grams). DDD = WHO defined daily dose (2023 ATC/DDD index). SD = standard deviation. Deviation (%) = (mean PDD/DDD − 1) × 100. Green = deviation > +5% (PDD exceeds DDD); red = deviation < −5% (PDD below DDD); grey = within ±5%. *Amox-Clav DDD reflects weighted mean of IV (3.0 g) and oral (1.5 g) forms; Ciprofloxacin reflects weighted mean of IV (0.8 g) and oral (1.0 g). ‡ n ≤ 2; interpret with caution. — = no prescriptions recorded in that month. The DDD is a statistical unit for drug utilisation comparison, not a recommended clinical dose.*
